# Social determinants of cognitive aging across 39 countries

**DOI:** 10.1101/2024.12.05.24318553

**Authors:** Victor Gilles, Syrine Salouhou, Rémi Vallée, Hugo Spiers, Michael Hornberger, Antoine Garnier-Crussard

## Abstract

Distinguishing between normal and pathological cognitive aging is challenging because there is no typical older person. The variation in age-related cognitive decline is not arbitrary. Several non-modifiable (e.g. genetic) and potentially modifiable (e.g. high blood pressure, smoking, and hearing loss) risk factors are associated with faster cognitive aging. Beyond these individual-level risk factors, a growing body of evidence has identified associations between cognitive impairment and social, economic, and environmental factors. These associations are even more pronounced in developing countries, notably because of greater disparities in education and socioeconomic status. We examine the association between demographic, social, economic, environmental wellbeing and spatial cognitive aging among 593,746 participants in 39 countries. Cognition is assessed using Sea Hero Quest, a spatial navigation video game that predicts spatial ability in the real world. The social, economic, and environmental well-being of older adults, as measured by the Global AgeWatch Index (GAWI), is negatively associated with age-related decline in spatial ability. In particular, the GAWI Health and Environment subscores are strongly correlated with the effect of age on wayfinding performance. We also found that gender differences in spatial navigation skills increase with age, and even more so in countries with greater gender inequality, as estimated by the Gender Inequality Index. Our results show that spatial cognitive aging must be understood as a dynamic, heterogeneous process that is strongly linked to potentially modifiable environmental and social factors.

## Introduction

Population aging represents a unique demographic shift occurring globally for the first time. By 2030, one in six people worldwide will be aged 60 or older, and by 2050, the number of people in this age group will have doubled to 2.1 billion. The number of people aged 80 and older is expected to triple between 2020 and 2050, reaching 426 millions [1]. This demographic shift is accompanied by a rise in age-related diseases: in 2017, the burden of age-related diseases represented 51.3% of the total global disease burden [2].

Dementia is one of the most prominent age-related diseases, marked by a considerable decline in cognitive abilities, which leads to dependence in daily activities [3]. Normal cognitive aging also involves a gradual cognitive decline. Distinguishing between normal and pathological cognitive aging is essential to optimize patient medical care, particularly for early identification of the disease before major neuronal damage occurs. Pathophysiological biomarkers (e.g. PET, CSF or blood-based biomarkers) of neurodegenerative diseases may enable in vivo characterization of pathological processes, distinguishing ‘normal’ ageing-related cognitive changes from early neurodegeneration. Blood biomarkers of AD are promising low-cost diagnostic tools [4], yet their accuracy vastly depends on the cognitive phenotype of the tested population [5]. The use of blood biomarkers needs to be paired with comprehensive cognitive phenotyping in clinical practice to avoid misdiagnosis due to false positives. Cognitive phenotyping is challenging because there is no typical older person. Some individuals in their eighties have cognitive abilities similar to many 30-year-olds, while others experience a significant cognitive decline much younger.

The differences in age-related cognitive decline are not arbitrary. Several factors are associated with cognitive aging, including non-modifiable genetic predispositions and potentially modifiable risk factors such as hypertension, smoking, and hearing loss [6, 7]. Beyond these risk factors at the individual-level, a growing body of evidence has identified associations between cognitive disorders and social, economic and environmental factors [8–13]. For instance social isolation is a risk factor for dementia [14, 15], and social inequalities involving age, gender, ethnicity, purchasing power and overall health lead to major disparities in the development, diagnosis and treatment of cognitive disorders [16–25]. These inequalities are not only more frequent but also more pronounced in developing countries shaped by the interactions of lower education and socioeconomic status, a higher cardiovascular disease burden, and genetic variability [26, 27]. These within-country inequalities increase the proportion of people experiencing adverse life conditions, thereby accelerating their cognitive ageing. Most modifiable risk and protective factors for cognitive decline and dementia demonstrate sex/gender differences in their rate and/or risk expression, with these differences often being more pronounced in developing countries [28, 29] or in populations with adverse socioeconomic conditions [30]. For instance, country-level gender inequalities are associated with brain structural differences [31], and in a previous study we also showed them to be associated with gender differences in spatial ability performance [32].

Although population aging began in high-income countries, such as Japan where 30% of the population is over 60, it is now low- and middle-income countries that are experiencing the greatest shift in age distribution. By 2050, over 65% of the world’s population over 60 years will live in low- and middle-income countries. The combined impact of this demographic shift and these environmental risk factors underscores the importance of studying the effects of socioeconomic and environmental factors on cognitive aging, particularly in populations beyond the WEIRD (Western, Educated, Industrialized, Rich and Democratic) countries that are usually studied in scientific research [33]. A citizen of a WEIRD country is 37 times more likely to appear in a medical study from top medical journals than a citizen of a non-WEIRD country [34]. In recognition of these challenges, the World Health Organization is leading the United Nations Decade of Healthy Ageing (2021–2030) [35]. This initiative is designed to improve the well-being of older adults, their families, and their communities. Investigating the determinants of cognitive ageing is thus closely aligned with these major international priorities.

Studies of cognitive ageing at the population level across several countries are rare, and even rarer are those that include participants from low- and middle-income countries. Most previous studies focused on the cultural determinants of cognition in the average population, i.e. did not study cognitive ageing per se. Quesque *et al.*, showed that up to 23% of variance in social cognition scores can be explained by cultural differences [36], while Coutrot *et al.* showed a positive correlation between country-level spatial navigation score and the Gross Domestic Product per capita [32]. However, none of these studies reported differences in cognitive aging trajectory. The socioeconomic determinants of a country’s average cognitive score can differ greatly from those of its rate of cognitive ageing. Another set of studies did focus on the structural determinants of ageing — particularly in South America, but mainly through the prism of brain correlates [37–40]. A Magnetic Resonance Imaging (MRI) study across 15 countries showed that the discrepancies between brain age and chronological age were associated with structural socioeconomic inequality, especially in Latin American and Caribbean countries [41]. Vidaurre *et al.* used MRI brain images across 52 countries and found that the association between female-male brain differences and gender inequality increases with age [42]. However, these differences at the brain level do not necessarily convert to differences in cognition or clinical status. For instance,

Ravndal *et al.* showed that sex differences in age-related brain decline are unlikely to explain the higher AD diagnosis prevalence in women [43]. Thus, a cross-country assessment of the societal determinants of cognitive ageing is needed to complement these neuroimaging results and inform the health policies that governments will need to develop, as well as the normative benchmarks that clinicians will need to use when handling the impending surge in age-related neurocognitive diseases.

In this study, we will investigate the association between demographic, social, economic and environmental well-being factors and cognitive aging among 715,295 participants in 46 countries. Spatial cognitive aging will be assessed by comparing the spatial abilities of younger (20–30 years old) and older (50–65 years old) participants playing Sea Hero Quest, a spatial navigation video game that is predictive of spatial ability in the real world [44, 45]. We use spatial navigation as a proxy for cognition. Spatial navigation is particularly interesting because it involves many cognitive processes, including interpreting a map, planning a multi-stop route, remembering the route, monitoring progress along the route and updating the route plan, and transforming the bird’s eye view into an egocentric perspective [46]. It relies on different brain structures such as the hippocampus (episodic memory, map representations, place cells), the retrosplenial cortex (egocentric and allocentric translation), or the posterior cingulate cortex (landmark position and attention). All of these brain structures are affected by Alzheimer’s disease in preclinical and prodromal stages [47], and are therefore often implicated in age-related cognitive decline. However, other cognitive functions are not directly probed by our spatial navigation task, such as purely memory-based or executive functions, which are also often affected by age-related cognitive decline. Spatial navigation is also interesting because it is a non-verbal cognitive function, suitable for cross-country – and therefore cross-language – experiments. The Sea Hero Quest project has gathered one of the largest and most diverse behavioral dataset, including participants from every country in the world and 150 000 participants above 50 y.o [48].

## Results

Sea Hero Quest is a spatial navigation video game available on tablets and smartphones. It was designed to quantify the player’s sense of direction through a series of tasks. Players were given the possibility to enter demographic information such as their age, gender, and nationality. The first two levels were tutorial levels and did not require any spatial ability. They were designed to familiarize the participants with the game controls, and incidentally test their motor skills. Here we focus on wayfinding levels, where the player is asked to memorize a map of an aquatic environment with a set of ordered checkpoint 1. When the player is ready, the map disappears and they have to navigate as quickly as possible to the checkpoints, in the correct orders. The first two levels are tutorial levels as no sense of direction is required. To quantify spatial ability, we used the length of the trajectories in different levels: the shorter the trajectory, the better the spatial ability.

Similarly to previously published studies based on the Sea Hero Quest dataset, to provide a reliable estimate of spatial navigation ability we only examined data from participants who completed at least 11 levels of the game (including the first 4 wayfinding levels: levels 6, 7, 8 and 11), and who entered their age, gender and nationality [49]. The focus of the current study being the differences in cognitive aging across countries, we only included participants from countries with at least 100 participants between 50 and 75 years old. This left us with 715,295 participants across 46 countries (320,876 females, mean age = 36.78 years, SD = 15.10 years). Figure 1 shows that after a continuous decline with age (i.e. an increase in trajectory length), spatial performance starts to re-increase after 78 y.o. Performance at tutorial levels follow the same trends, but with a much weaker effect of age. To quantify these differences, we computed the effect size of age on motor and spatial ability. We used Hedge’s g to contrast younger (20-30 y.o.) and older (50-65 y.o.) participants. We found *g* = 0.94 (95% CI = [0.93, 0.95]) for spatial ability, and *g* = 0.23 (95% CI = [0.22, 0.24]) for motor ability. We previously interpreted the increase in performance after a certain age as a strong selection bias, causing the performance of older participants to be substantially higher than would be expected in unselected participants of the same age [32, 49]. For instance, ‘superagers’ participants may be more likely to play SHQ and have higher-than-expected cognitive abilities. This pattern also exists for training performance, but to a much smaller scale. As shown in Supplementary Figure 1, this selection bias age varies between countries, from 48 y.o. in Uganda to 88 y.o in Finland. To avoid biasing our aging estimates with this phenomenon, we decided to exclude countries where the selection bias age is under 65 y.o. We also excluded participants above 65 y.o. in the remaining countries, leading to a final dataset of 593,746 participants across 39 countries (268,708 females, mean age = 35.70 y.o., SD = 13.38 y.o.), see Table S1 and Figure 2. We defined a younger group with participants between 20 and 30 y.o. (N = 289,762, 43% females) and an older group with participants between 50 and 65 y.o. (N = 118,132, 53% females). 50 to 65 years old can seem a bit young to form an ‘older participants’ group, which often starts at 65 y.o., but on top of mitigating the selection bias, this makes sense in the context of Alzheimer’s research. Spatial navigation impairment in considered as one of the earliest behavioral change in AD, and can occur decades before the onset of other cognitive symptoms [47, 50]. Since the focus of SHQ is on the early diagnosis of AD, studying ‘young older adults’ is relevant.

**Fig. 1.**
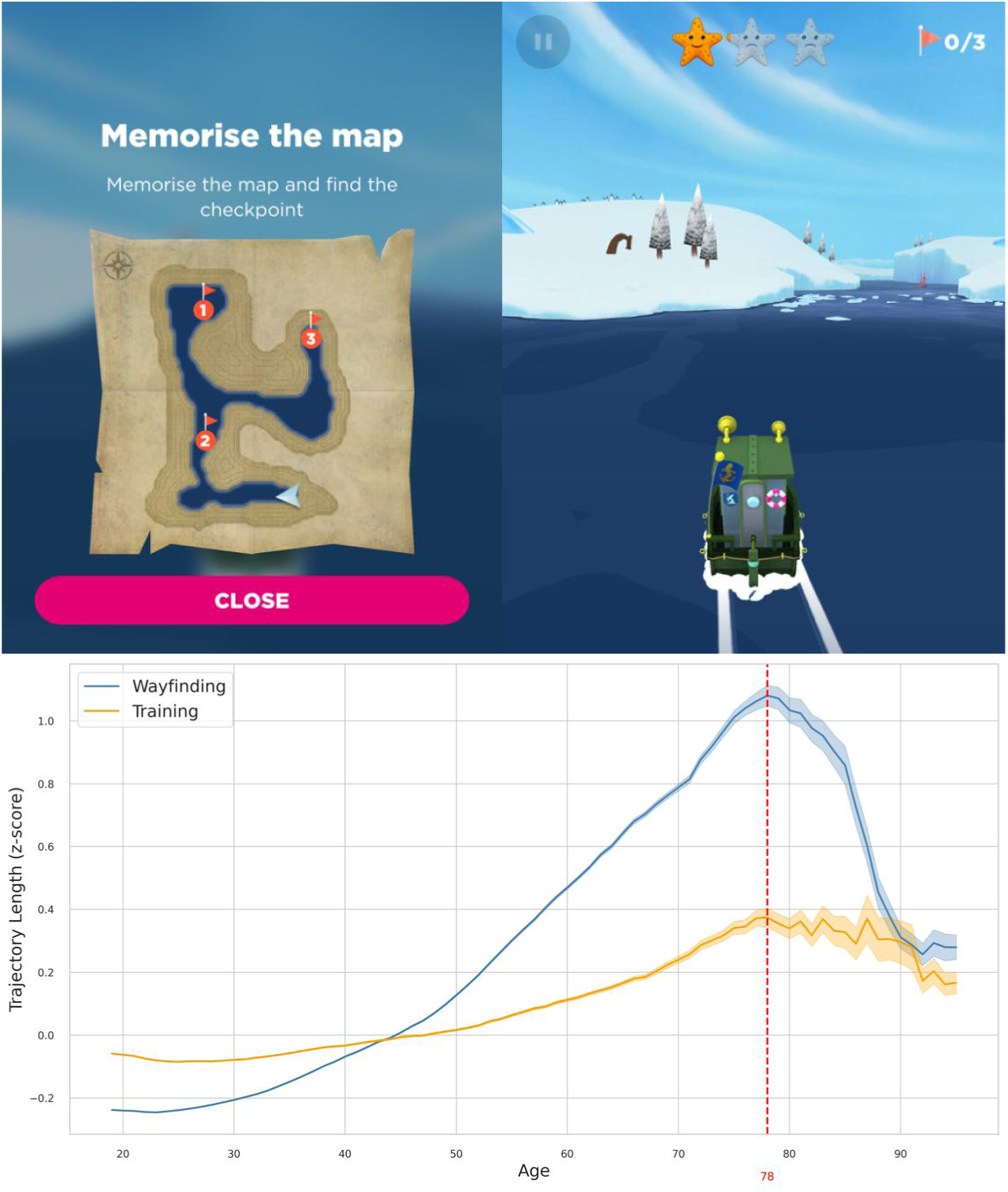
Sea Hero Quest level and trajectory length. | Top: Sea Hero Quest screenshots representing the map and a navigation scene for wayfinding level 11. Bottom: Trajectory length as a function of age for wayfinding (in blue) and training (in yellow) levels. Trajectory length is inversely proportional to performance. Data points represent the average trajectory lengths within five-year windows. The red vertical bar indicates the global maximum of the curves, and the starting age of the selection bias. Error bars correspond to standard errors.

**Fig. 2.**
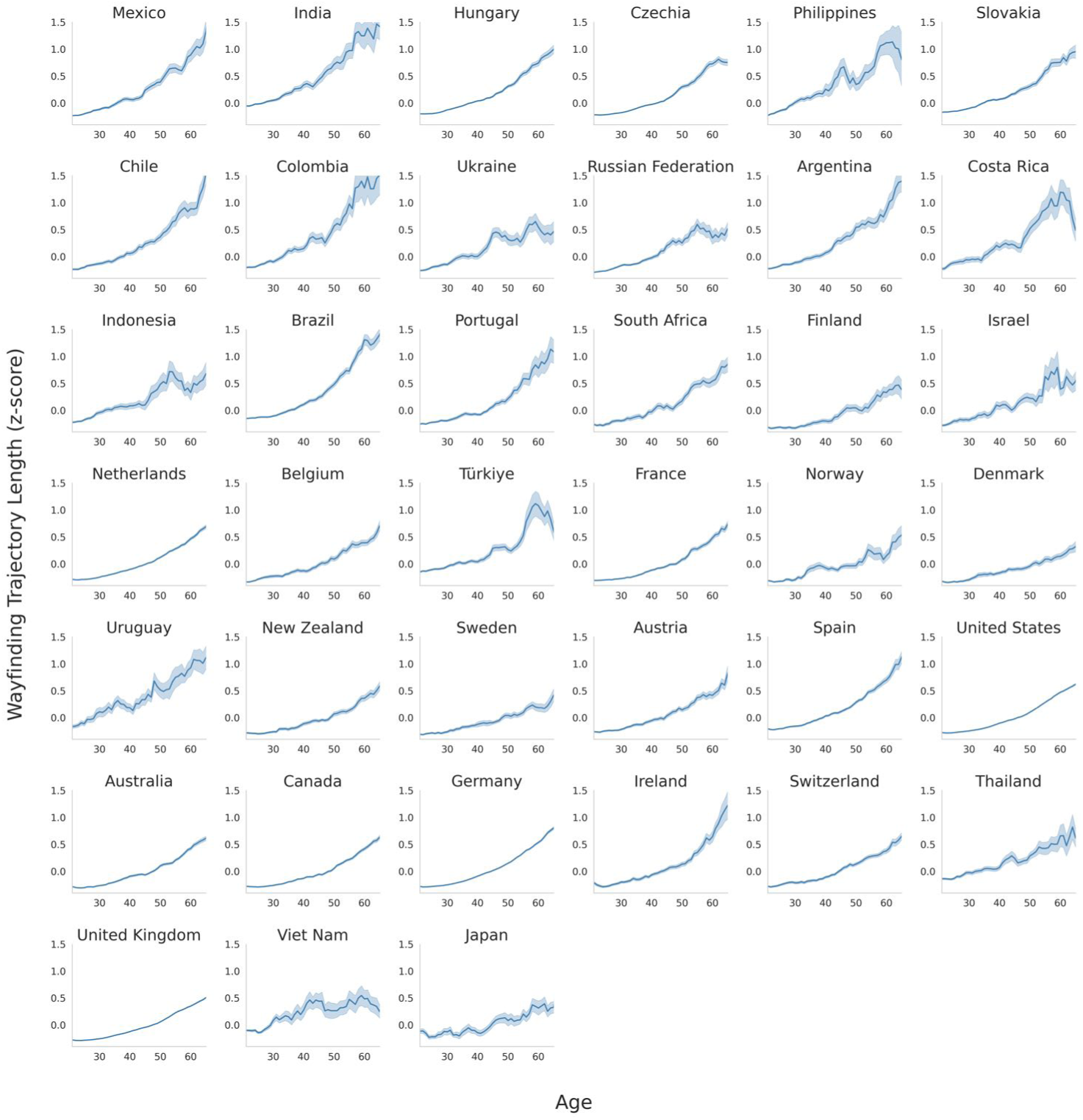
Wayfinding trajectory length as a function of age in 39 countries. | Z-scored wayfinding trajectory length from 20 to 65 y.o. Data points represent the average z-scored trajectory lengths within five-year windows. See Figure S1 for age range up to 95 y.o, and Figure S2 for the effect sizes between the older group (50-65 y.o.) and the younger group (20-30 y.o.). The countries are ranked in order of decreasing age effect size, see also Figure S2. Error bars correspond to standard errors. Standard errors are inversely proportional to the sample size, so larger error bars in developing countries are likely caused by lower sample sizes.

### Spatial cognitive aging and socio-economic indicators

We observed a similar pattern of spatial cognitive decline with age across all countries (Figure 2). However, the rate of this decline presents stark differences between countries. As previously we used Hedge’s g to contrast younger (20-30 y.o.) and older (50-65 y.o.) participants, see Figure 4. Hedge’s g ranged from 0.59 in Japan to 1.76 in Mexico, positive values indicating longer trajectories for older participants.

To understand the association between these cross-country ageing differences and their socio-economic status, we used several indicators: the GDP per capita, the life expectancy, the Human Development Index (HDI), the Global AgeWatch Index (GAWI), the Gini coefficient, and the Gender Inequality Index (GII), see Methods for details about these indicators. Jointly using these indices in a linear model to predict cognitive ageing is challenging are they obviously all are strongly correlated (e.g. Pearson’s correlation between GDP per capita and Life Expectancy is r = 0.70, between GDP per capita and HDI is r = 0.81). To overcome this collinearity challenge, we used the Least Absolute Shrinkage and Selection Operator (LASSO) regression. This regularization technique penalizes less significant predictors, producing a more parsimonious and interpretable model. The LASSO identifies the most relevant predictors by showing only the variables with substantial contributions to the outcome. We run two LASSO regressions: the first one with the outcome being the effect size of age on wayfinding trajectory length, the second one with the outcome being the average wayfinding trajectory length in each country. In both cases, the predictors are the country-level indices. As shown in Figure 3, the first LASSO regression identifies the GDP per Capita and the GAWI as the most relevant predictors corresponding to the minimum cross-validated mean squared error (MSE). The second LASSO regression identifies the Gini coefficient as the most relevant predictor for the mean trajectory length. The fact that the GAWI is selected by the first LASSO regression but not by the second one makes sense, as it is the only indicator focussing on age-related inequalities. In the following, we choose to focus on this indicator. As this study focuses on country-level differences in cognitive ageing, we decided to examine its association with the age-specific indicator GAWI rather than the broader indicator GDP per capita. However, one should keep in mind that the two are strongly correlated (Pearson’s *r* = 0.87, *p <* 0.001).

**Fig. 3.**
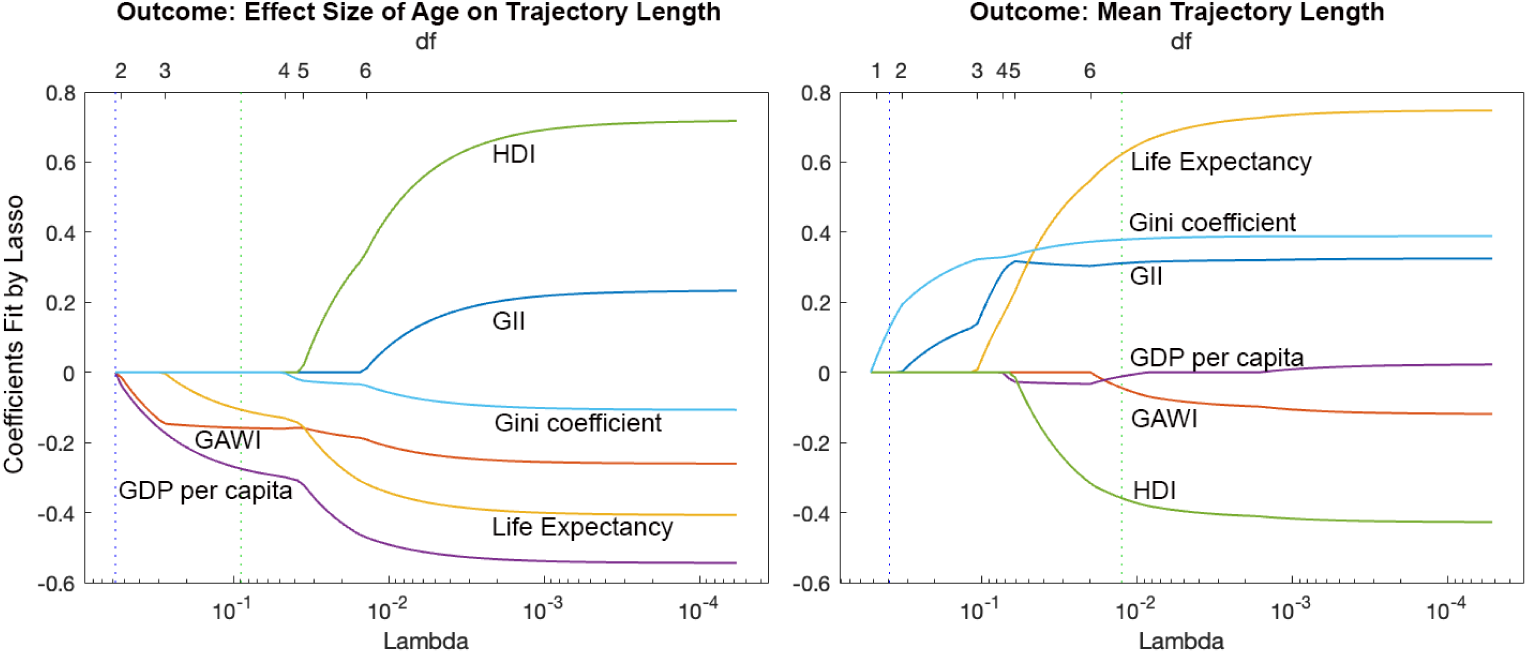
Trace Plots of LASSO Coefficients. | Left: the outcome is the effect size of age on wayfinding trajectory length in each country. Right: the outcome is the average wayfinding performance in each country. The lower x-axis represents the L1 penalization *Lambda*: the larger Lambda (i.e. the more to the left), the sparser the model. The upper x-axis contains the degrees of freedom (df), meaning the number of nonzero coefficients. The green dashed line indicates the value of Lambda with a minimum cross-validated mean squared error (MSE). The blue dashed line indicates the greatest Lambda that is within one standard error of the minimum MSE.

### Cognitive aging and the Global AgeWatch Index

To better understand what are socio-economic inequalities associated with aging across the globe, the Global AgeWatch Index (GAWI) was developed in 2013 [51]. The most recent iteration, updated in 2015 [52], employs a four-category classification system, including Income Security, Health Status, Capability, and Enabling Environment (see Methods). We found a strong negative correlation between GAWI and the effect of age on wayfinding ability (Pearson’s *r* = −0.57, *p <* 0.001). Both Health and Environment GAWI subscores are strongly negatively correlated with the effect of age on wayfinding ability (Pearson’s *r* = −0.59 and *r* = −0.60 respectively, *p <* 0.001), see Figure 4. The Capability subscore was moderately correlated with the age effect (*r* = −0.33, *p* = 0.04) and the Income subscore was not significantly correlated with the age effect (*r* = −0.22, *p* = 0.17). To control whether these differences were not related to difference in age or gender distribution across countries, we computed a linear mixed model with trajectory length as the dependent variable, age, gender and their interaction as fixed effects, and age, gender and their interaction as random effects clustered by countries (see Methods). As with the Hedge’s g, we obtained a strong correlation between the GAWI and the age slopes clustered by countries (*r* = −0.56, *p <* 0.001).

**Fig. 4.**
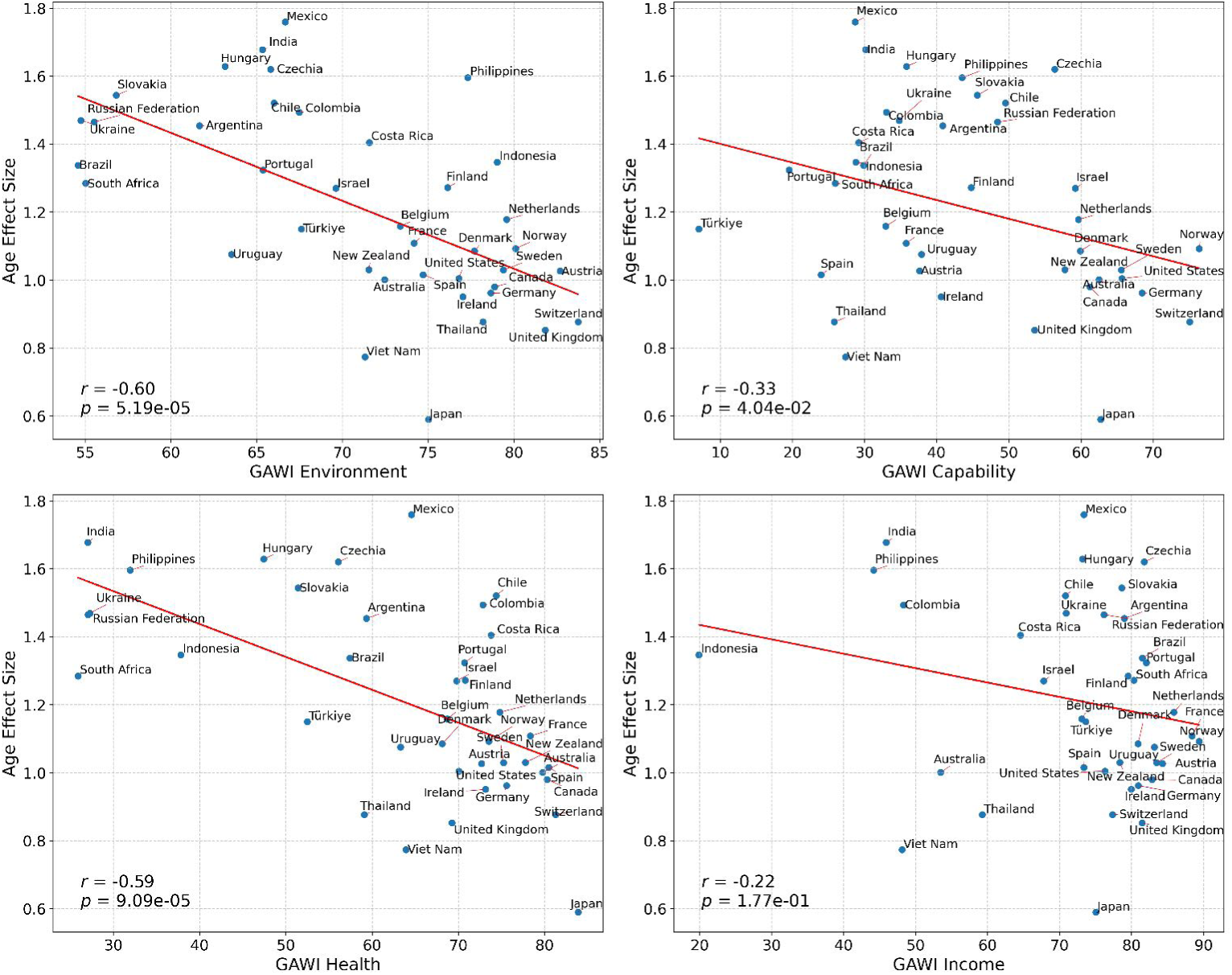
Association between Age Effect Size and GAWI subscores. | Positive age effect sizes indicate better wayfinding performance for younger participants. Larger GAWI values indicate better living condition for older participants. The red lines illustrate the strong negative correlations between the Health (*r* = −0.59, *p <* 0.001) and Environment (*r* = −0.60, *p <* 0.001) GAWI subscores and the age effect size.

To study the association between cognitive ageing and the GAWI beyond the age of 65, we used the Hedge’s g to contrast younger (20-30 y.o., N=253,529) and older (50-98 y.o., N=157,409) participants with no upper age limit, in all 46 countries. We did not find a significant correlation between the global GAWI score and the effect of age (*r* = −0.26, *p* = 0.07), see Fig. S2a. As mentioned earlier, this negative result is likely due to the fact that the selection bias mitigates the rate of cognitive decline after a certain age. For instance, countries with early selection bias age such as Albania, Serbia and Uganda have a smaller effect of age because their older participants are impacted by selection bias earlier than participants from other countries. To mitigate this bias while including participants beyond 65 years old, we excluded countries with an age of selection bias earlier than 75 y.o. and capped the older group at 75 y.o. This left us with 22 countries, 127,777 participants in the younger group and 130,662 in the older group. In this population, despite the smaller number of included countries, we found a significant correlation between the global GAWI score and the effect of age (*r* = −0.48, *p* = 0.02), see Fig. S2b. The fact that older and younger groups are more balanced in countries with a later age of selection bias is consistent with this bias being partly associated with older generations more easily adopting mobile and video games. To quantify this trend, we found a positive correlation between the cross-country differences in selection bias age observed in Fig. S1 and the GAWI global score (*r* = 0.46, *p* = 0.001), see Fig. S3. This means that countries with stronger socio-economic inequalities associated with aging have earlier selection bias age.

### Association with the level of education

350,314 of the included participants entered their education level among 4 options: “no formal’ (N = 8,595), ‘high-school’ (N = 89,371), ‘college’ (N = 100,248) or ‘university’ (N = 152100). We merged ‘university’ and ‘college’ into a unique ‘tertiary education’ level (72%, N = 252,348). This was notably motivated by the fact that ‘university’ and ‘college’ have different meanings in different countries. For instance, the word ‘college’ can refer to a community college, technical school, or liberal arts college in some countries, or used interchangeably with ‘university’ in others. Similarly, we merged ‘high-school’ and ‘no formal’ into a unique ‘secondary education and lower’ level (28%, N = 97,966). We chose not to analyse separately the ‘no formal’ group because its relative low sample size makes it more liable to selection bias. Figure S3 represents the 4 dimensions of the GAWI as a function of the effect size of age for the participants for received secondary education or lower (top) and tertiary education (bottom). The associations between the Health and Environment GAWI subscores and the effect of age on wayfinding ability are strengthened in the ‘secondary education or lower’ population (*r* = −0.54*, p <* 0.001 for Environment, *r* = −0.53*, p <* 0.001 for Health) compared to the ‘tertiary education’ population (*r* = −0.36*, p* = 0.02 for Environment, *r* = −0.46*, p* = 0.002 for Health). These results indicate that the level of education modulates but do not override country-level socioeconomic indicators.

### Sex difference in spatial cognitive aging

In Figure 5 we show the trajectory length as a function of age for males and females in each country. On average, males have better wayfinding performance than females. We showed in [32] that the gender effect size was correlated with a country-level metric of gender inequality: countries having a high level of gender inequality in their society also have a larger gender difference in Sea Hero Quest performance. Here we are interested in the difference in the spatial ability slopes across lifespan between male and female participants in the 39 included countries. As in the previous section, we computed a linear mixed model with trajectory length as the dependent variable, age, gender and their interaction as fixed effects, and age, gender and their interaction as random effects clustered by countries. We computed Pearson’s correlation between the interaction coefficient clustered by countries and the Gender Inequality Index (GII). The GII is a country-level index for the measurement of gender inequality taking into account reproductive health, empowerment, and labor market participation [53]. We found a negative correlation (Figure 6, *r* = −0.63*, p <* 0.001), indicating that in countries with higher gender inequality, females have a faster cognitive decline in terms of wayfinding abilities compared to men.

**Fig. 5.**
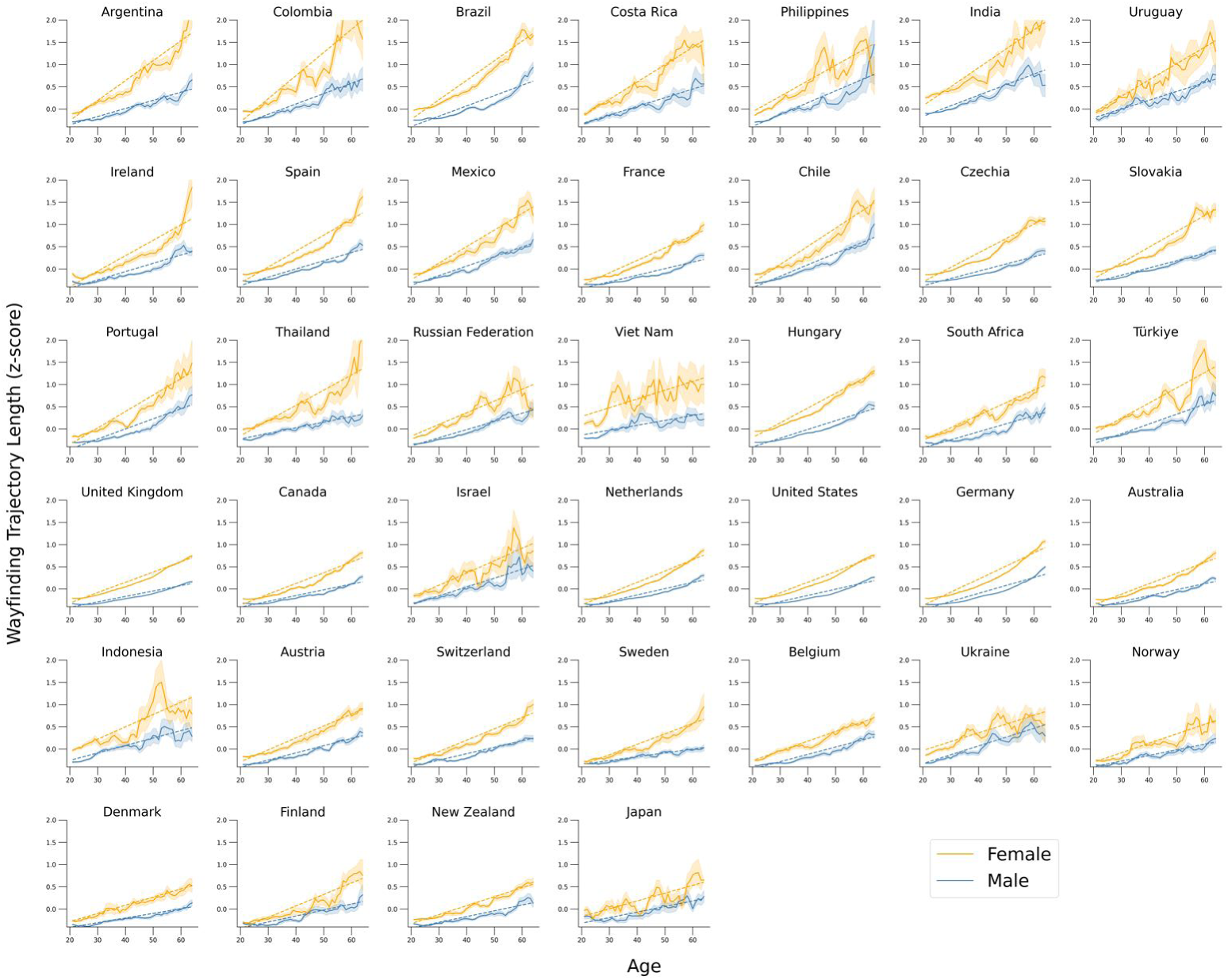
Wayfinding trajectory length as a function of age, stratified by gender, in 39 countries. | Countries are sorted according to the value of the age x gender interaction term clustered by country in a linear mixed model. Data points represent the average trajectory lengths within five-year windows. Error bars correspond to standard errors.

**Fig. 6.**
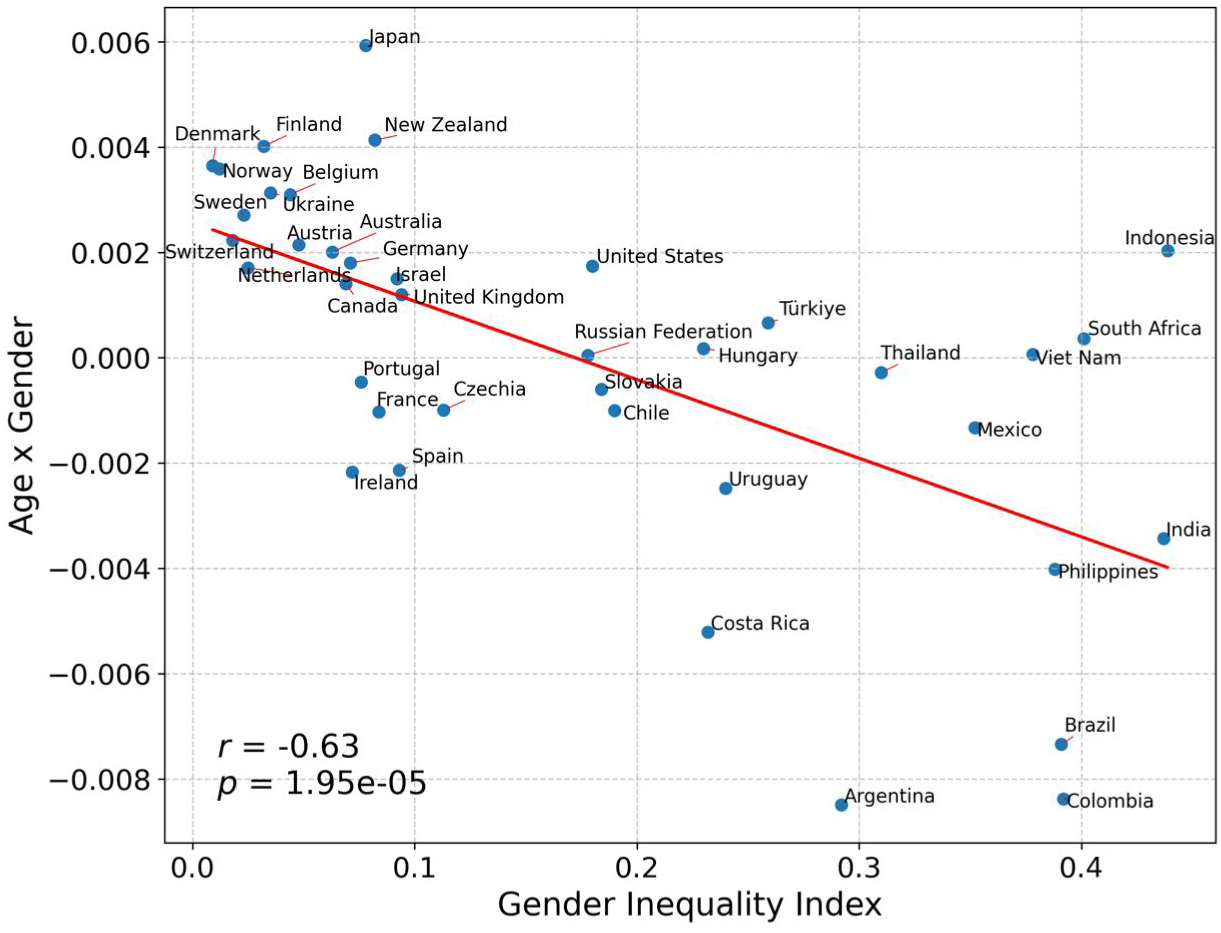
Interaction between Age and Gender as a function of the Gender Inequality Index (GGI) | The Age x Gender interaction terms are calculated by a linear mixed model, with Trajectory Length as the response variable, Age, Gender and their interaction as fixed effects, and Age, Gender and their interaction as random effects clustered by countries. Negative Age x Gender interaction values correspond to gender differences increasing with age. Increasing GGI values correspond to larger gender differences within countries. We found a significant negative correlation (*r* = −0.63, *p <* 0.001), indicating that in countries with greater gender inequality, females experience a more rapid cognitive decline than males.

## Discussion

In this study, we investigate how age-related cognitive decline assessed using a spatial navigation video game varies in over 590 000 participants from 39 countries. Using LASSO regression to predict spatial cognitive ageing with different country-level socio-economic indicators, we show that the Global AgeWatch Index (GAWI) is one of the most relevant predictors. We find that the social, economic and environmental well-being in older adults as measured by the GAWI, is negatively associated with age-related decline in spatial ability. In particular, the Health and Environment GAWI subscores are strongly correlated with the effect of age on wayfinding performance. We also found that gender differences in spatial navigation skills increase with age, and even more so in countries with greater gender inequality, as estimated by the Gender Inequality Index. Our results show that the dynamics of cognitive aging vary considerably with socio-economic indicators at the country level. This is particularly true for health (life expectancy, psychological well-being) and environmental (social connections, physical safety, civic freedom, access to public transport) indicators. On the contrary, indicators of income (pension, poverty rate in older age, Gross National Income per capita) and capability (employment and educational status of older people) indicators are less associated with country differences in cognitive decline. These findings are consistent with previous work that have shown how environmental characteristics can affect individual and global health [10, 54–57]. For instance, more socially active older adults experience less cognitive decline in old age [58], and loneliness increase the risk for all-cause dementia by 30%, even when controlling for other modifiable risk factors [59]. From a mechanistic perspective, the link between cognition and social activity can be explained by increased engagement in interpersonal interactions. This enhances cognitive reserve, providing individuals with meaningful social roles and a sense of purpose that can have neurohormonal effects on the brain [58]. Higher levels of physical activity, which are potentially linked to the ‘enabling environment’ GAWI subscore, have been associated with slower cognitive decline during aging. This has been observed even among individuals with preclinical Alzheimer’s disease, for whom physical activity has been linked to reduced tau accumulation [60]. Socioeconomic status is linked to disparities in brain health. In particular, higher socioeconomic status in adults has been associated with slower cognitive decline, greater brain volume, and reduced white matter hyperintensity burden – a biomarker of cerebrovascular damage [61]. Access to healthcare, which can be indirectly captured by the GAWI, is also associated with lower risk of cognitive decline [62]. These results support the need for public policy [63], such as age-friendly transportation (e.g. free public transport) could be used to improve cognitive health in oder adults by promoting a less sedentary and more social behavior [64, 65].

While gender differences in some cognitive tasks have been shown to correlate with national indicators of gender equality [32, 66], the interaction between age and gender has rarely been analyzed (although see [24]). Our results suggest that the decline in spatial navigation skills is more pronounced in women over the course of the adult life. The gender difference in the slope of spatial cognitive decline is correlated with the Gender Inequality Index (GII), indicating that in countries with lower gender inequality, males and females show smaller differences in cognitive aging. These differences can be interpreted in three ways. First, the acceleration of spatial cognitive decline in women could be explained by a cumulative effect: the longer females live in an unequal environment, the greater the contrast with men. Second, women born in countries with high levels of gender inequality probably had reduced opportunity in childhood and early adulthood, contributing to accelerated cognitive decline later in life. Longitudinal studies have shown that early-life exposure explain a significant amount of the variance in cognitive functioning in later adulthood [67]. Finally, spatial cognitive gender differences could be greater for people in their sixties than in their twenties because they have not been exposed to the same inequalities, which tend to decrease with time [68]. In this way, differences between men and women at a particular age could be a snapshot of gender inequalities experienced by that generation. The age-related decline in spatial navigation skills being more pronounced in women can also be interpreted in the light of sex differences in dementia prevalence. Previous studies showed that AD is more prevalent in women at most ages, while it is not the case for other types of dementia, such as vascular dementia [69]. This is consistent with the fact that spatial navigation deficits are much more present in AD than in other dementia types [47], as mentioned in the introduction.

There are several limitations to our study. First, the demographic profile of our participants is limited, as we only have access to self-reported general information such as their age, gender, and country of residence. Access to more precise information such as their socio-professional categories, reading or intellectual game habits would allow us to estimate their cognitive reserve, which is often associated with cognitive decline trajectories [54, 70–73]. Although we asked participants about their gender, we only allowed binary response options (male/female). This limits how the variable can be used and could confuse it with biological sex. Knowing about participants’ country of birth or migration history would also be of interest since childhood environment significantly shapes adult cognition [49, 74]. Indeed, some participants may have experienced various socioeconomic conditions throughout their lives. This could result in exposure misclassification, leading to the dilution, introduction of heterogeneity, or partial conflation of the observed associations between country-level indicators and cognitive decline. Similarly, knowledge of participants’ neighborhood at a finer scale would be helpful, as previous research has shown that greater access to green spaces from childhood through to adulthood can help to slow down the rate of cognitive decline in later life [75, 76]. We did not directly select participants based on their cognitive health, and a few participants with dementia or mild cognitive impairment may have been included in the sample. However, we assume it is unlikely that participants with mild cognitive impairment or dementia would have voluntarily engaged with and completed 11 levels of our video game, without external support. Another limitation is the selection bias inherent in cross-sectional studies of unselected participants. A recent study showed that people with higher incomes and better computer skills are more likely to agree to participate in such scientific experiments [77]. As they get older, unselected participants are more likely to have above-average cognitive health, as shown by the inflection point in Figure 1. As our cognitive task is based on a video game, this selection bias is likely reinforced by the effect of digital exclusion, which particularly affects older people [78] and participants from less developed countries [79]. The positive correlation between selection bias age and GAWI (Fig. S3) shows that this between-country difference is at least partly driven by cultural and socio-economic factors, such as the adoption of technology among older people, its accessibility, their purchasing power... This explains that a culturally and socio-economically homogeneous group of countries such as Balkan and Eastern-European countries share similar selection bias ages (see Fig. S1). It would also be interesting to directly test whether there is a greater digital age gap in developing countries than in developed countries. This seems to be the case in this study, as Table S1 shows that there are more older participants in more developed countries. To mitigate this tendency, we only included participants from countries with at least 100 participants in the older group. Digital device use has been positively associated with cognitive reserve and cognition [72, 80], in addition to general health [81, 82] and social connection [83–85] in older people. However, two results mitigate these concerns. First, the effect of age on pure motor skills, as assessed during the tutorial levels, was four times smaller than the effect of age on wayfinding skills. This indicates that the effects measured with SHQ and presented in this manuscript have a strong cognitive component and are not simply a matter of familiarity with digital technology. Second, we have previously reported that SHQ performance is predictive of performance at a similar task in the real world [44]. This result was recently replicated in a cohort of older adults (54-74 years old), but not for SHQ levels identified as either too easy or too difficult, which were not included in the current analysis [45]. Selection bias is more problematic in countries where the inflexion point in cognitive performance occurs before 65 y.o., such as Uganda, Albania, Serbia or Greece… in these countries the Sea Hero Quest database is unable to provide an unbiased cognitive ageing estimate, and further data collection should be performed, for instance with representative longitudinal cohorts. This would allow to specifically include more older participants to mitigate the digital age gap in developing countries compared to developed countries. Collecting longitudinal data would also allow to make causal claims, while only correlations can be tested with our cross-sectional dataset. We have started to collect such data in the UK with the Understanding Society cohort [86], but it needs to be generalized to other – less WEIRD – countries. In particular, Africa should be more represented, as we only managed to include South Africa in the current analysis.

## Conclusions

Our study highlights the diversity of spatial cognitive aging in different countries across the five continents and their association with socioeconomic indicators and gender inequalities across the lifespan. The challenge of cognitive aging, with an exponential increase in cognitive impairment and dementia in the coming decades, is even more important to understand and prevent in countries of the global South, by addressing growing socioeconomic inequalities, in addition to traditional individual risk factors. Cognitive aging must be understood as a dynamic, heterogeneous process that is strongly linked to environmental and social factors. From a prevention point of view, a better understanding of the drivers of these inequalities is a key point for preparing specific interventions in education, global health and social connectedness. These interventions need to be implemented at the neighborhood level, but also led by strong public policies at the national level, in order to better prepare for the challenges of aging populations around the world.

## Methods

### Data

The design and the data collection process for SHQ have been thoroughly described in previous manuscripts [32, 49]. SHQ has initially been developed as a task that could benchmark navigation ability for cognitive assessment in dementia research. The project arose from a collaboration between a private company (Deutsche Telekom) who funded the development and the marketing of the app, professional game developers (Glitchers) and academics. Several strategies were taken to advertise for participants, from film adverts about the study on social media, to specific adverts sent to Deutsche Telekom’s customers. In a few weeks, SHQ became viral, and was even the most downloaded app on the Apple Store for a day. This snowball effect is what we attributed to being able to recruit 3.9 million participants drawn from every country in the world. The data has been collected between 2016 and 2019 with the mobile and tablet video game Sea Hero Quest, freely available on all app platforms.

### Informed consent and ethics approval

This study has been approved by the UCL Ethics Research Committee. The ethics project ID number is CPB/2013/015. Participants were made aware of the purpose of the game within the opening screen. Demographics such as their age, gender and home country were provided by consenting participants. They were asked whether they were willing to share their data with us and were guided as to where they could opt out. The opt-out was always available in the settings.

### Cognitive Task

In this study we used the wayfinding task in the video game Sea Hero Quest. At the beginning of each wayfinding level, participants were shown locations (checkpoints) to visit on a map. The map disappeared, and they had to navigate a boat through a virtual environment to find the different checkpoints. The checkpoints were typically not encountered in the order in which they were passed, but had to be navigated by returning from one checkpoint to another (Figure 1). The first two levels were tutorial levels to familiarize the participant with the game commands.

### Participants

A total of 3 881 449 participants played at least one level of the game; 60.8% of the participants provided basic demographics (age, gender, home country) and 27.6% provided more detailed demographics (home environment, level of education). For the gender, the specific question was “What gender are you?” (not sex), and the two options were “Male” and “Female”. To provide a reliable estimate of spatial navigation ability, we examined data only from participants who had completed a minimum of 11 levels of the game (including the first 4 wayfinding levels: levels 6, 7, 8 and 11) and who entered all their demographics. To test whether there were more ‘dropouts’ across the levels in the older group than in the younger group, we computed the number of younger and older players at each level (1, 2, 6, 7, 8, 11), divided by the number of players at level 1. For the younger group, these ratios respectively are 1, 0.97, 0.82, 0.77, 0.72, 0.58. For the older group, these ratios respectively are 1, 0.97, 0.76, 0.72, 0.67, 0.53. The dropout rate is slightly higher in the older group, suggesting that our results can be an underestimation of the true effects. The focus of the current study being the differences in cognitive aging across countries, we only included participants from countries with at least 100 participants between 50 and 75 years old. This left us with 715 295 participants across 46 countries (320876 females, mean age = 36.78 years, SD = 15.10 years). To avoid biasing our aging estimates with the selection bias (see in Results and Discussion), we decided to exclude participants from countries where the selection bias age is under 65 y.o. We also excluded participants above 65 y.o. in the remaining countries, leading to a final dataset of 593 746 participants across 39 countries (268 708 females, mean age = 35.70 y.o., SD = 13.38 y.o.), see Table S1.

### Wayfinding performance

We collected the trajectory of each participant across levels 1, 2, 6, 7, 8 and 11. The coordinates of participants’ trajectories were sampled at Fs = 2 Hz. As in previous studies [32, 49], we computed the trajectory length in pixels, defined as the sum of the Euclidean distance between the points of the trajectory. The first two levels only reflected video gaming skill (motor dexterity with the game controls) as no sense of direction was required to complete them. For each level, we took the z-score of participants’ trajectory length. This normalization leads to more interpretable results across levels, which all have different size. We defined the overall training trajectory length metric as the average between the z-scored trajectory lengths of training levels 1 and 2. We defined the overall wayfinding trajectory length metric as the average between the z-scored trajectory lengths of wayfinding levels 6, 7, 8, and 11. As this metric is based on the trajectory length, it varies as the opposite of the performance: the longer the trajectory length, the worse the performance.

### Age-related selection bias

The inflexion point corresponding to the start of the selection bias is different for each country (see Figure S1). To quantify it, we took the age corresponding to the global maximum of the curve representing the overall wayfinding trajectory length as a function of age in each country.

### Country-level indicators

To quantify the socio-economic status of the countries included in our analysis we used:

- **The Gini coefficient**, a measure of statistical dispersion intended to represent the income inequality, the wealth inequality, or the consumption inequality within a nation [87].
- **The Life Expectancy**, as estimated in the World Population Prospects database by the United Nations Population Division [88].
- **The GDP per capita**, as estimated by the International Monetary Fund’s World Economic Outlook Database [89]
- **The Human Development Index**, a statistical composite index of life expectancy, education and per capita income indicators [90], gathered by the United Nations Population Division [91].

For each indicator we used the values collected the closest to 2017, which was the median date of the Sea Hero Quest data collection (data collected between 2016 and 2019, with a peak in 2017).

To quantify socio-economic inequalities associated with aging across the globe, the Global AgeWatch Index (GAWI) was developed in 2013 [51]. The most recent iteration, updated in 2015 [52], employs a four-category classification system, including:

- **Income security**: pension income coverage, poverty rate in old age, relative welfare of older people, GNI per capita
- **Health status**: life expectancy at 60, Healthy life expectancy at 60, psychological well-being
- **Capability**: employment of older people, educational status of older people
- **Enabling environment**: social connections, physical safety, civic freedom, access to public transport.

Country-level gender inequality has been quantified with the **Gender Inequality Index (GII)**, an indicator developed by the United Nations Development Programme available for more than 170 countries. [92, 93]. We used the latest available GII values (2022). This index is a composite measure to quantify the loss of achievement within a country due to gender inequality. It uses three dimensions to measure opportunity cost: reproductive health, empowerment, and labor market participation. This indicator has recently been used in a study comparing brain health across countries [29, 41].

## Statistical Analysis

### - LMM computation

We computed a linear mixed model with trajectory length as the dependent variable, age, gender and their interaction as fixed effects, and age, gender and their interaction as random effects clustered by countries:

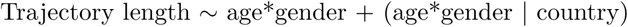

To study the effect of age across countries we used the age random slopes clustered by countries. To study the sex differences in cognitive aging across countries we used the age x gender random interaction values clustered by countries The parameters of the linear mixed models were estimated with the maximum likelihood method, and the covariance matrix of the random effects were estimated with the Cholesky parameterization.

### - Hedge’s g

Hedge’s g between group 1 and group 2 is defined as:

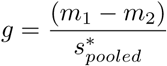

where *m_i_*is the mean of group *i*, and *s*^∗^ is the pooled and weighted standard deviation:

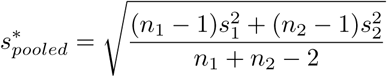

with *n_i_*the sample size of group *i*, and *s_i_* the standard deviation of group *i*.

The 95% confidence intervals displayed in this manuscript are exact analytical confidence intervals based on iterative determination of noncentrality parameters of noncentral t or F distributions

### - LASSO regression

The Least Absolute Shrinkage and Selection Operator (LASSO) regression is similar to a standard regression, but it penalizes the number of predictors, leading to a sparser and more interpretable model [94]. The formula for the LASSO regression is as follows:

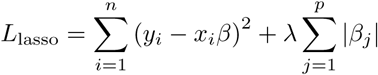

Where *β* are the coefficients (i.e., the importance) of the selected indicators x in predicting the outcome, and *λ* penalizes the number of variables(the higher *λ*, the sparser the model). We run two LASSO regressions with two different outcomes: the first one with the effect size of age on the overall wayfinding trajectory length, the second one on the average overall wayfinding trajectory length, for each country. In both cases the predictors are the country-level indices, i.e. the GDP per capita, the life expectancy, the Gini coefficient, the Human Development Index, the Global AgeWatch Index, and the Gender Inequality Index.

## Code and Data availability

The code and data necessary to reproduce the results presented in this manuscript are available at https://osf.io/e6tgk/?viewonly=1890283b477b4f259f75e8a7c9117045. We also set up a portal where researchers can invite a targeted group of participants to play SHQ and generate data about their spatial navigation capabilities. Those invited to play the game will be sent a unique participant key, generated by the SHQ system according to the criteria and requirements of a specific project. https://dash.seahero.quest/wiki/ Access to the portal will be granted for non-commercial purposes.

## Data Availability

The code and data necessary to reproduce the results presented in this manuscript are available at https://osf.io/e6tgk/?view_only=1890283b477b4f259f75e8a7c9117045

## Supplementary Material

**Fig. S1.**
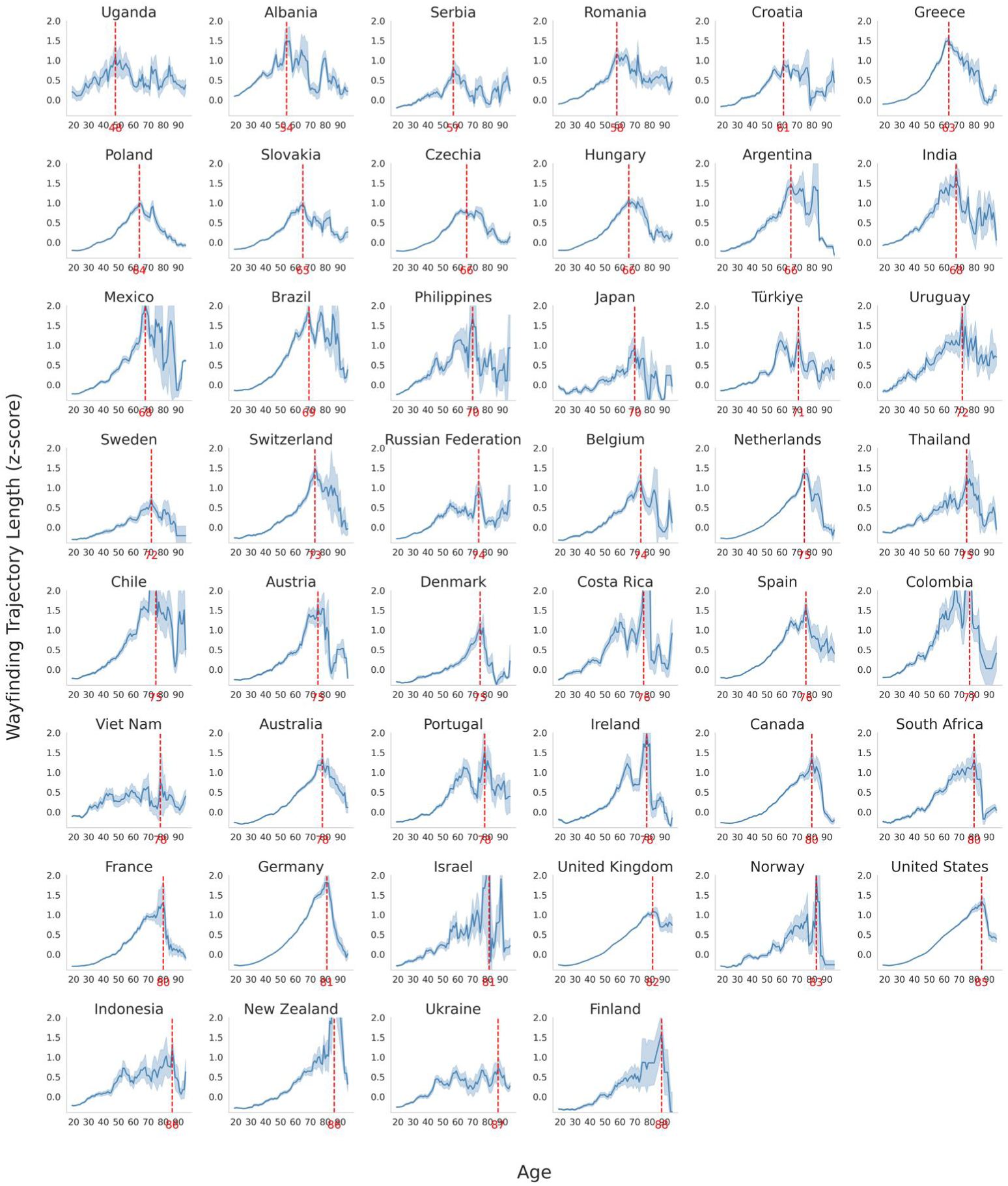
Wayfinding trajectory length as a function of age, in 46 countries. — Z-scored wayfinding trajectory length from 20 to 95 years. The red vertical line represents the global maximum of each country curve. Countries are sorted according to this value. Data points represent the average trajectory lengths within five-year windows. Error bars correspond to standard errors.

**Fig. S2.**
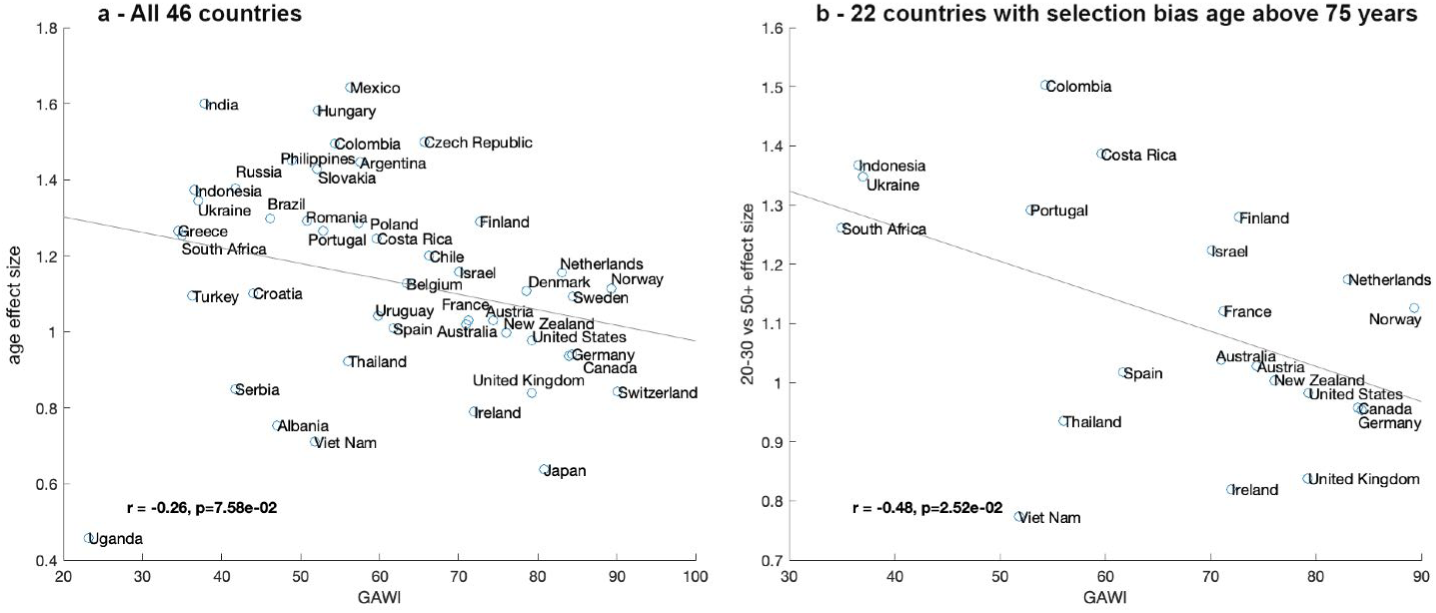
Association between the rate of cognitive decline and age-related socio-economic inequality (GAWI) — **a**, In all 46 countries and no upper age limit. **b**, In the 22 countries where the age of selection bias was above 75 years.

**Fig. S3.**
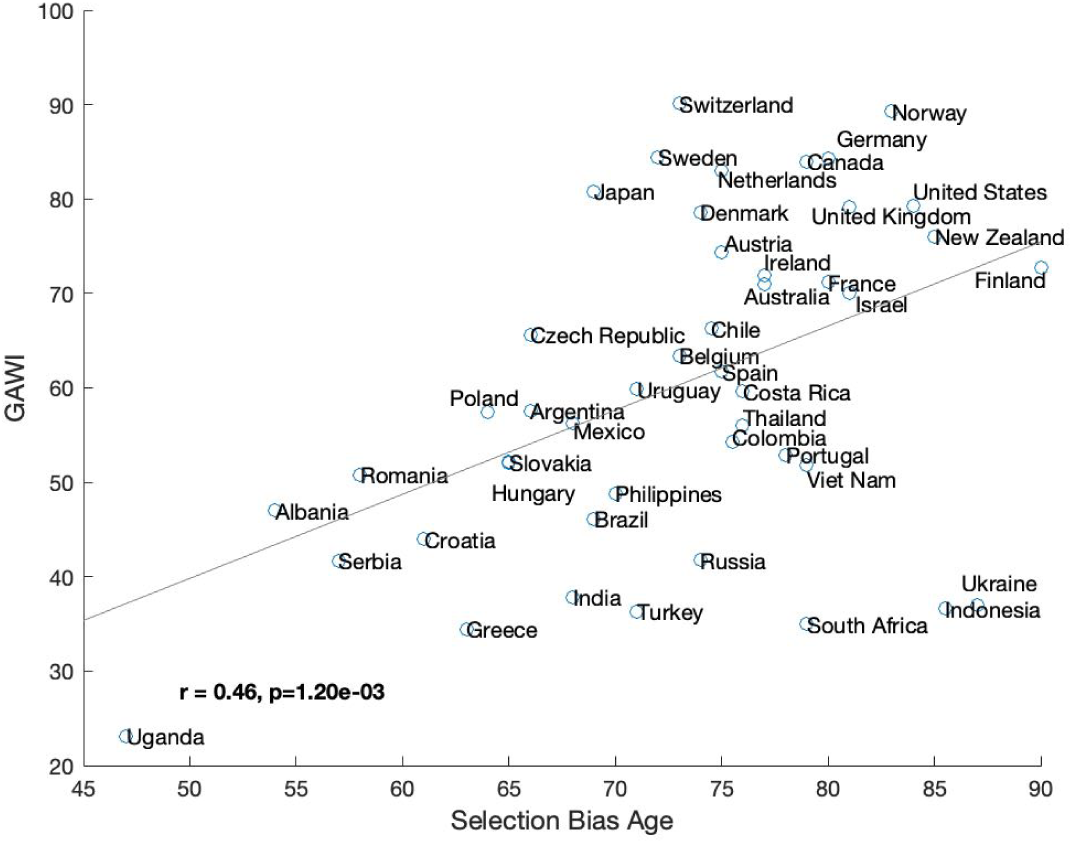
Association between selection bias age and age-related socio-economic inequality (GAWI), in 46 countries. — We found a positive correlation between these variables (*r* = 0.46*, p* = 0.001), meaning that the stronger the inequalities, the earlier the selection bias occurs.

**Fig. S4.**
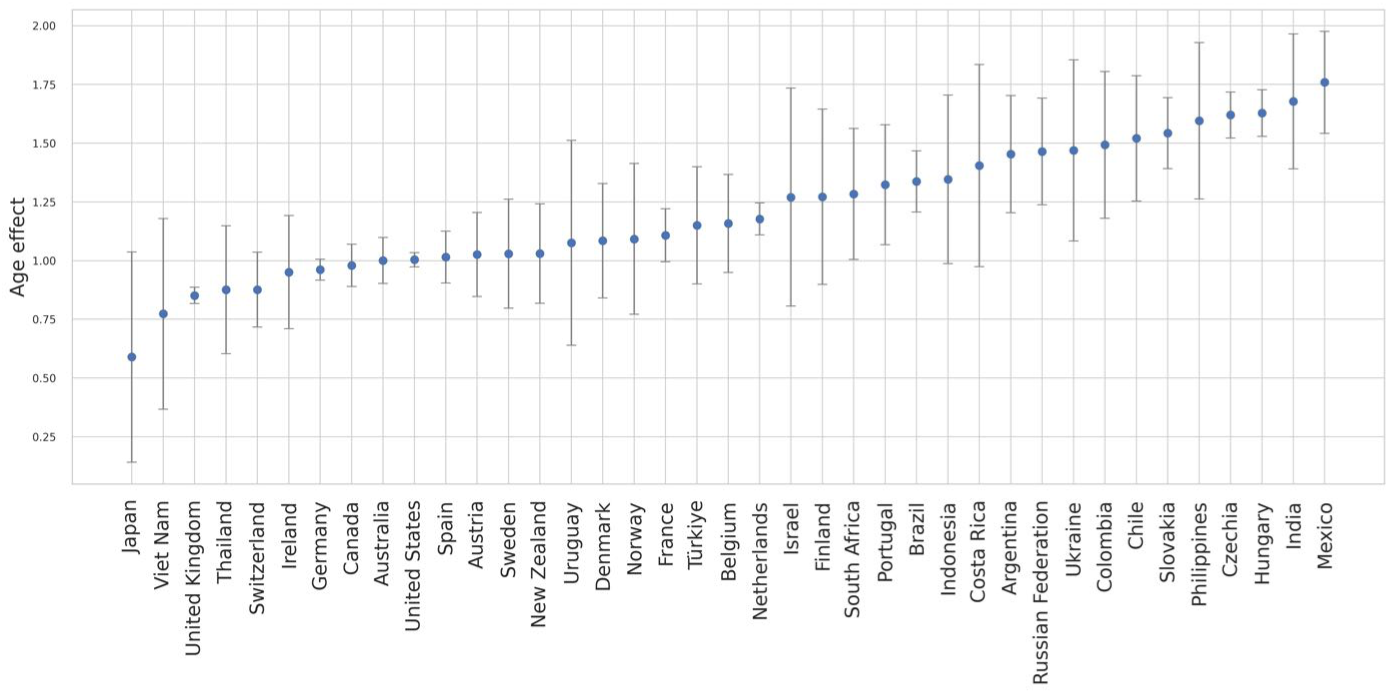
Effect size of age on wayfinding trajectory length in 39 countries. | The effect size is the Hedge’s g between the older group (50-65 y.o.) and the younger group (20-30 y.o.). Positive values indicate shorter wayfinding trajectories for younger participants. Error bars correspond to 95% Confidence Intervals.

**Fig. S5.**
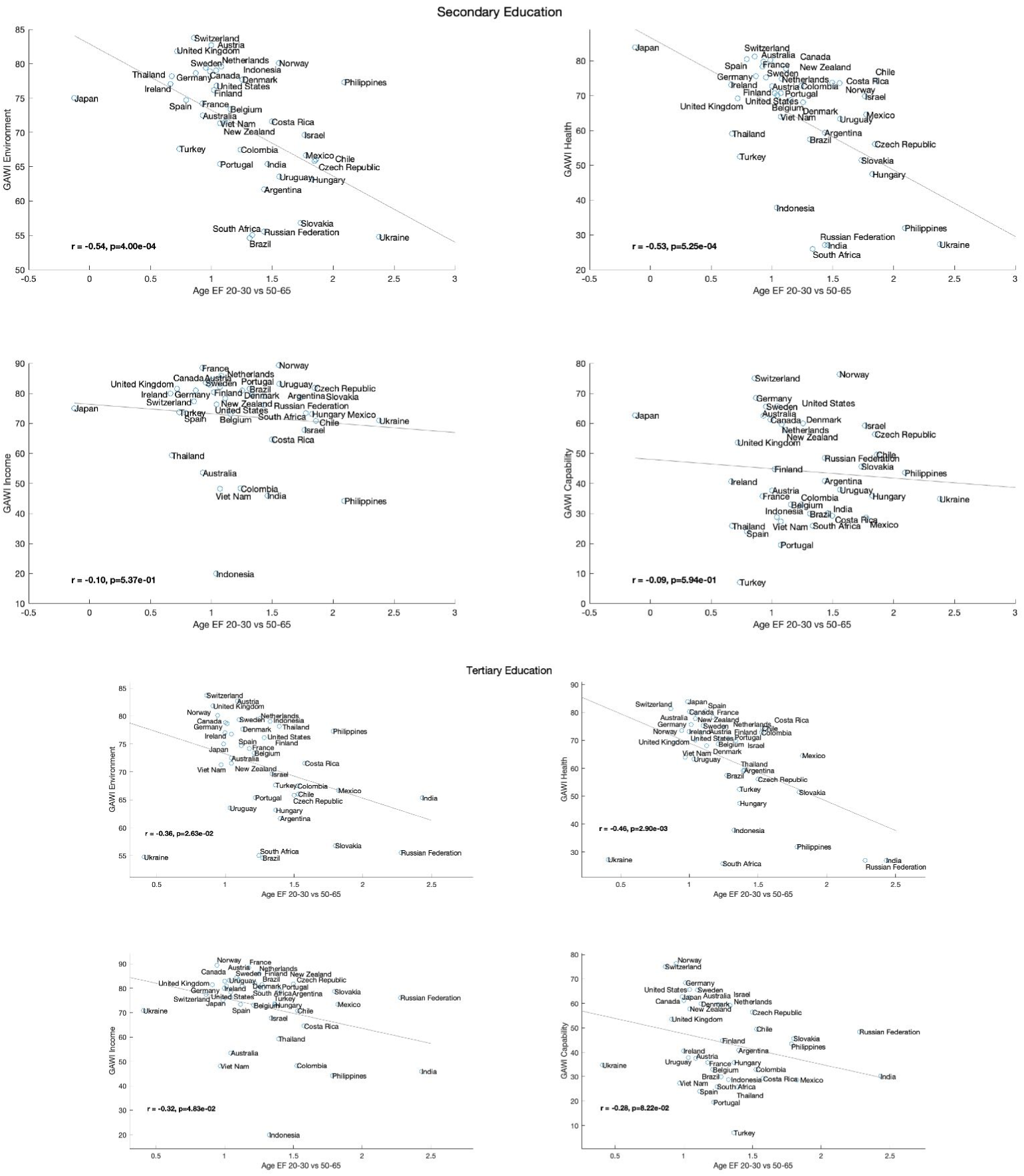
Association between Age Effect Size and GAWI subscores for participant with secondary (top) or tertiary (bottom) education. | The effect size (Age EF in the x-axis) is the Hedge’s g between the older group (50-65 y.o.) and the younger group (20-30 y.o.). Positive age effect sizes indicate better wayfinding performance for younger participants. Larger GAWI values indicate better living condition for older participants.

**Table S1.** For each country, number of participants included in the analysis (N), their mean age and standard deviation, the proportion of females, and the proportion of participants between 50 y.o. and 65 y.o.

| Country | World Region | N | Age ($M \pm SD$) | Females | >50 y.o. |
| --- | --- | --- | --- | --- | --- |
| Argentina | America | 3151 | 31.79 ± 12.02 | 0.38 | 0.12 |
| Australia | Oceania | 13932 | 37.48 ± 13.86 | 0.51 | 0.24 |
| Austria | Europe | 4457 | 35.31 ± 12.96 | 0.43 | 0.19 |
| Belgium | Europe | 3075 | 36.39 ± 13.90 | 0.40 | 0.23 |
| Brazil | America | 11283 | 31.46 ± 12.05 | 0.43 | 0.12 |
| Canada | America | 16094 | 37.72 ± 14.00 | 0.49 | 0.25 |
| Chile | America | 2690 | 32.54 ± 12.29 | 0.39 | 0.13 |
| Colombia | America | 2433 | 30.50 ± 11.14 | 0.40 | 0.09 |
| Costa Rica | America | 963 | 33.30 ± 12.28 | 0.43 | 0.14 |
| Czech Republic | Europe | 39264 | 29.01 ± 9.92 | 0.43 | 0.05 |
| Denmark | Europe | 2713 | 34.58 ± 12.84 | 0.37 | 0.16 |
| Finland | Europe | 1373 | 32.99 ± 11.96 | 0.41 | 0.12 |
| France | Europe | 11821 | 34.78 ± 13.11 | 0.40 | 0.18 |
| Germany | Europe | 63412 | 39.20 ± 13.17 | 0.46 | 0.27 |
| Hungary | Europe | 42519 | 29.04 ± 9.93 | 0.41 | 0.04 |
| India | Asia | 7536 | 26.33 ± 8.03 | 0.17 | 0.03 |
| Indonesia | Asia | 3237 | 27.23 ± 9.14 | 0.28 | 0.04 |
| Ireland | Europe | 2602 | 37.97 ± 12.77 | 0.41 | 0.21 |
| Israel | Middle East | 999 | 32.60 ± 11.84 | 0.26 | 0.10 |
| Japan | Asia | 686 | 36.19 ± 12.82 | 0.36 | 0.18 |
| Mexico | America | 6456 | 28.79 ± 10.56 | 0.37 | 0.07 |
| Netherlands | Europe | 31644 | 35.49 ± 13.05 | 0.53 | 0.19 |
| Norway | Europe | 2006 | 32.33 ± 11.83 | 0.42 | 0.10 |
| New Zealand | Oceania | 3010 | 37.35 ± 13.57 | 0.53 | 0.23 |
| Philippines | Asia | 3205 | 27.59 ± 9.63 | 0.46 | 0.05 |
| Portugal | Europe | 3597 | 33.44 ± 10.99 | 0.39 | 0.09 |
| Russia | Asia | 8268 | 28.06 ± 8.98 | 0.34 | 0.04 |
| Slovakia | Europe | 18581 | 28.42 ± 9.57 | 0.42 | 0.04 |
| South Africa | Africa | 2098 | 35.39 ± 12.91 | 0.42 | 0.18 |
| Spain | Europe | 12598 | 35.71 ± 12.56 | 0.39 | 0.17 |
| Sweden | Europe | 3591 | 32.11 ± 12.10 | 0.39 | 0.11 |
| Switzerland | Europe | 5050 | 38.95 ± 13.62 | 0.43 | 0.27 |
| Thailand | Asia | 3221 | 29.44 ± 10.71 | 0.40 | 0.08 |
| Turkey | Middle East | 4783 | 30.56 ± 10.05 | 0.33 | 0.06 |
| Ukraine | Europe | 2229 | 28.41 ± 9.65 | 0.32 | 0.06 |
| United Kingdom | Europe | 106354 | 41.20 ± 13.37 | 0.46 | 0.32 |
| United States | America | 139817 | 37.45 ± 13.80 | 0.51 | 0.24 |
| Uruguay | America | 724 | 35.74 ± 13.35 | 0.38 | 0.20 |
| Viet Nam | Asia | 2274 | 25.72 ± 9.01 | 0.31 | 0.05 |

